# Macular vessel density in diabetes and diabetic retinopathy with swept-source optical coherence tomography angiography

**DOI:** 10.1101/2020.02.19.20024174

**Authors:** Naiqiang Xie, Yan Tan, Sen Liu, Yining Xie, Shaoshuai Shuai, Wei Wang, Wenyong Huang

**Author notes:** Co-first authors (Naiqiang Xie, Yan Tan). **Corresponding author:** Wenyong Huang, MD&PhD, Professor of Ophthalmology, Vice director of Department of Preventive Ophthalmology, Zhongshan Ophthalmic Center, State Key Laboratory of Ophthalmology, Sun Yat-sen University, 54S.Xianlie Road, Guangzhou, China 510060, Wei Wang, MD&PhD, Zhongshan Ophthalmic Center, State Key Laboratory of Ophthalmology, Sun Yat-sen University, 54S.Xianlie Road, Guangzhou, China 510060. **Acknowledgments:** None. **Funding:** None.

## Abstract

**Purpose:** Previous studies on the association between macular vessel density (VD) and diabetic retinopathy had conflicting conclusions. This study assessed the alterations of macular VD, as well as other factors, in diabetic patients using swept-source optical coherence tomography angiography (SS-OCTA) in a large-scale sample from Chinese communities.

**Methods:** Patients with type 2 diabetes without history of ocular treatment were recruited from 2017 to 2018. The average and quadrant parafoveal vessel density (PVD) were obtained with a commercial SS-OCTA device (Triton, Topcan, Japan). Univariate and multivariate linear regression was used to analyse the correlation of PVD with diabetic retinopathy (DR), diabetic macular edema (DME), HbA1c, and other factors.

**Results:** A total of 919 patients were included in the final statistical analysis. After adjusting for other confounding factors, the DR patients had significantly lower average PVD (β= −1.062, 95% CI = −1.424 to −0.699, P < 0.001) in comparison with those without DR. In addition, the patients with mild DR or vision-threatening diabetic retinopathy (VTDR) also had significantly lower PVD (P < 0.001 for mild DR, and P = 0.008 for VTDR) compared with those without DR. Age and HbA1c were also significantly related to PVD measurements, as shown by multivariable linear regression. Participants with DME had a significantly lower average PVD and temporal PVD than those without DME (P < 0.05).

**Conclusions:** Reduced PVD was independently associated with more severe DR, older age, higher HbA1c level, and the presence of DME. These findings provide manifest evidence to suggest that macular vessel alterations play a role in the pathogenesis of DR.

## Introduction

Diabetes retinopathy (DR), a common issue in patients with diabetes, is one of the major causes of blindness in developed countries.[1] DR features vascular alteration in different layers of the retina, which may lead to visual impairment. The pathogenesis of DR remains unclear, and recent studies suggested that the alteration of retinal microvasculature, especially in the macula, might contribute to the presence and progression of DR.[2, 3] Optical coherence tomography angiography (OCTA), a non-invasive fundus angiography method that can show the morphology of retinal microvasculature, become widely used to assess DR.[4] One of the most studied parameters of OCTA was vessel density (VD), which is implicated in the presence and progression of DR.[5, 6]

Several studies evaluated the VD changes in DR; however, the relationship between VD and DR remains controversial. Some studies have found that non-DR patients of diabetes mellitus (DM) exhibit similarities to non-DM patients with superficial retinal VD, but other studies have found that both the superficial and deep capillaries of diabetic patients have decreased VD.[7-10] Moreover, some studies have found that the correlation between VD and DR in superficial capillaries exceeds that of deep retinal VD, superficial retinal perfusion density, or foveal avascular zone (FAZ) area.[11, 12] The relationship between VD and DME is unclear: some studies have found that DME’s VD declines while others found no difference.[13, 14] The relationship between VD and the glycaemic index has conflicting conclusions: some studies have found that VD is negatively correlated with systemic factors—such as fasting blood glucose, postprandial blood glucose, and HbA1c—but other studies have found that VD is not associated with HbA1c or the duration of the disease.[15-17]

The discrepancy of these findings may be related to several factors, including inadequately adjusting for confounding factors, such as age, gender, blood pressure, serum lipids, ocular treatment (PRP and anti-VEGF); insufficient technology, such as non-SS-OCTA and manual-measurement PVD; and small sample size.[18] In addition, we are unaware of any SS-OCTA studies of diabetes patients from Chinese communities. Therefore, the objective of this study was to assess the alterations of macular VD in diabetic patients and related factors using SS-OCTA.

## Materials and Methods

### Participants

This cross-sectional study was performed at the Zhongshan Ophthalmic Centre (ZOC), affiliated with Sun Yat-sen University in China. The Ethics Committee at ZOC approved the study protocol. The study was performed according to the tenets of the Helsinki Declaration and we obtained signed informed consent forms from all participants. Patients with type 2 diabetes, who were aged 40 years and over and without history of ocular treatment, were recruited from the community health system in Guangzhou, China. The participants were excluded if they had a history of systemic diseases, except for diabetes, such as malignant tumours, ischemic heart disease, stroke, cancer, or kidney disease; a history of heart bypass, thrombolysis, or kidney transplantation; cognitive impairments or mental illnesses that made them unable to complete questionnaires or cooperate with examiners; or any eye diseases, other than DR, such as glaucoma, vitreous macular disease (vitreous haemorrhage or retinal detachment), or amblyopia. They were also excluded if they had a history of intraocular procedures, such as retinal laser or intraocular injection history, glaucoma surgery, cataract surgery, laser myopia surgery, abnormal refractive mediums, corneal ulcer, pterygium, corneal opacity, poor fixation that caused poor quality of ocular imaging, spherical equivalent (SE) < −9.0D, astigmatism > +4.0 D, axial length (AL) > 26.0 mm, or a best corrected visual acuity (BCVA) of less than 20/200.

### General information and laboratory parameters

The demographic and systemic information, including age, sex, duration of diabetes, and medical history, were obtained using customized questionnaires and the information was confirmed after further inspection of outpatient history. The height, weight, systolic blood pressure (SBP), and diastolic blood pressure (DBP) were measured using standardized processes. The haemoglobin A1c (HbA1c), total cholesterol, triglyceride, and C-reactive protein levels were determined in a laboratory certified by the Chinese government.

### Ocular examination and SS-OCTA imaging

All participants underwent comprehensive ocular examinations, including slit lamp bioscopy, ophthalmoscopy, intraocular pressure (IOP), visual acuity, refraction, ocular biometry, retinal photography, and OCTA imaging. The optical low-coherence reflectometry (Lenstar LS900; Haag-Streit AG, Koeniz, Switzerland) was used for measuring ocular biometric parameters, including central corneal thickness (CCT), anterior chamber depth, lens thickness, and AL. Standardised 7-field colour retinal images adhering to Early Treatment Diabetic Retinopathy Study (ETDRS) criteria were obtained using a digital fundus camera (Canon CR-2, Tokyo, Japan) after full pupil dilation. Trained personnel graded the retinal images according to the guidelines of the United Kingdom National Diabetic Eye Screening Programme. DR severity was graded as R0, R1, R2, or R3. DME was graded as M0 (no maculopathy) or M1 (exudate within 1-disc diameter of the fovea, or a collection of exudates within the macula).

A commercial SS-OCTA instrument (DRI OCT-2 Triton; Topcon, Tokyo, Japan) was used for retinal microvasculature imaging, which had a central wavelength of 1050 nm with an axial resolution of 8 μm, a transverse resolution of 20 μm, and a speed of 100,000 A-scans per second. The parafoveal VD (PVD) measurements of the superficial capillary plexus were calculated automatically using the built-in software (version 2014.2.0.65). The PVD was defined as the proportion of the vessel area showing blood flow relative to the total area measured. The quadrant PVD (superior, inferior, nasal, and temporal) was rendered using an ETDRS grid overlay comprising the two inner rings. All OCTA images were acquired by a well-trained examiner. Images with low signal strength index (SSI < 50), one or more blinking artefacts, motion or double artefact (caused by poor fixation), a blurry rendition of the vascular system (caused by the non-transparency of the medium), the destruction of the retina’s anatomical characteristics (caused by the change of the macula), or slab errors were excluded.

### Statistical analyses

The Kolmogorov-Smirnov test was used to verify the normal distribution, and Fisher’s exact test was used for categorical variables. When normality was confirmed, a t-test was conducted to evaluate the inter-group difference of demographic, systemic and ocular parameters. Linear regression analysis was performed to compare PVD measurements with demographic or ocular parameters. The univariate analysis showed that the predictive variables were significant. These were then entered into the final multivariate equation using the stepwise method. A *p* value <0.05 was considered statistically significant. All analyses were performed using Stata Version (Stata Corporation, College Station, TX, USA).

## Results

### Demographic and clinical features of the participants

A total of 919 eyes (919 patients) were included in this study. Demographic and clinical characteristics of study participants are shown in Table 1. Among them, 381 (41.46%) patients were male, with a mean age of 64.8±7.1 years and an average duration of diabetes of 8.8±6.8 years. Of the 755 patients without DR, 300 (39.74%) were male, with a mean age of 64.8±7.1 years. Of the 104 patients with mild DR, 49 (47.12%) were male, with a mean age of 65.5±7.6 years. Of the 60 patients with vision-threatening diabetic retinopathy (VTDR), 32 (53.33%) were male, with a mean age of 63.6±6.5 years. Parameters relating to the use of insulin, duration of diabetes, SBP, BCVA, AL and retinal thickness differed among the groups (p < 0.05). The other parameters had no significant difference.

**Table 1.** Demographic and clinical characteristics of study participants.

| Characteristics | All | Non-DR | Mild DR | VTDR | P-value |
| --- | --- | --- | --- | --- | --- |
| No. of subjects | 919 | 755 | 104 | 60 | - |
| Gender | 381(41.46%) | 300(39.74%) | 49(47.12%) | 32(53.33%) | 0.056 |
| Use of insulin | 160(17.64%) | 103(13.84%) | 31(30.1%) | 26(43.33%) | **<0.001** |
| Mean age, year | 64.8±7.1 | 64.8±7.1 | 65.5±7.6 | 63.6±6.5 | 0.225 |
| Duration of diabetes, year | 8.8±6.8 | 8.2±6.5 | 11.2±7.7 | 12.2±7.4 | **<0.001** |
| HbA1c, % | 7.0±1.4 | 6.8±1.2 | 7.7±1.7 | 8.3±1.8 | **<0.001** |
| Height, cm | 160.2±8.1 | 160.0±8.1 | 160.6±8.3 | 161.2±7.3 | 0.465 |
| Weight, kg | 63.4±11.6 | 63.6±11.8 | 63.5±10.7 | 62.0±11.5 | 0.590 |
| Systolic blood pressure, mm Hg | 136.5±18.9 | 135.6±18.4 | 138.3±20.2 | 145.2±20.4 | **<0.001** |
| Diastolic blood pressure, mm Hg | 71.3±10.3 | 71.3±10.2 | 71.5±10.0 | 72.2±11.6 | 0.772 |
| Total cholesterol, mmol/L | 4.8±1.1 | 4.8±1.1 | 4.7±1.1 | 5.0±1.2 | 0.230 |
| Triglycerides, mmol/L | 2.2±1.5 | 2.2±1.5 | 2.1±1.3 | 2.4±2.3 | 0.519 |
| C-reactive protein, mg/L | 2.7±6.1 | 2.8±6.2 | 2.9±6.6 | 2.0±2.0 | 0.621 |
| BCVA, logMAR | 0.2±0.1 | 0.2±0.1 | 0.2±0.1 | 0.3±0.2 | **<0.001** |
| IOP, mmHg | 16.1±2.7 | 16.1±2.8 | 16.2±2.5 | 16.2±2.4 | 0.951 |
| CCT, µm | 546.5±30.3 | 545.9±30.9 | 548.1±29.1 | 551.5±25.1 | 0.328 |
| Axial length, mm | 23.4±0.9 | 23.5±0.9 | 23.4±0.9 | 23.2±0.9 | **0.021** |
| Cornea diameter, mm | 11.6±0.4 | 11.6±0.4 | 11.7±0.4 | 11.5±0.4 | 0.070 |
| Choroidal thickness, µm | 188.2±73.5 | 187.0±73.9 | 192.7±71.9 | 195.0±71.6 | 0.603 |
| Retinal thickness, µm | 275.9±19.5 | 274.9±16.3 | 273.9±13.1 | 290.0±43.0 | **<0.001** |

DR=diabetic retinopathy; VTDR= vision threatening diabetic retinopathy; BCVA=best corrected visual acuity; IOP= intraocular pressure; CCT=central corneal thickness. Bold indicates statistically significant.

### Association between parafoveal vessel density and DR

Figure 1 shows examples of OCTA images of non-DR and DR patients. The PVD difference is statistically significant among three groups. The PVD decreased as DR worsened, as shown in Figure 2. Univariate linear regression showed significant negative correlation between average PVD and DR status (Table 2). The results of further stepwise multivariable linear regression are shown in Table 3.

**Table 2.** Univariable linear regression analysis of average para-foveal vessel density.

| Univariate | Coefficient | 95%CI |  | P-value |
| --- | --- | --- | --- | --- |
|  |  | Lower | Upper |  |
| Mean age, year | -0.019 | -0.038 | -0.0001 | **0.049** |
| Gender (Male vs Female) | -0.327 | -0.601 | -0.053 | **0.020** |
| Duration of diabetes, year | -0.021 | -0.042 | -0.001 | **0.039** |
| Use of insulin (yes vs no) | -0.408 | -0.765 | -0.050 | **0.025** |
| HbA1c, % | -0.275 | -0.372 | -0.177 | **<0.001** |
| Height, cm | -0.010 | -0.027 | 0.007 | 0.242 |
| Weight, kg | -0.005 | -0.017 | 0.007 | 0.393 |
| Systolic blood pressure, mm Hg | -0.004 | -0.011 | 0.003 | 0.259 |
| Diastolic blood pressure, mm Hg | -0.003 | -0.017 | 0.010 | 0.606 |
| Total cholesterol, mmol/L | 0.018 | -0.109 | 0.145 | 0.779 |
| Triglycerides, mmol/L | 0.076 | -0.013 | 0.166 | 0.095 |
| C-reactive protein, mg/L | 0.004 | -0.019 | 0.026 | 0.747 |
| BCVA, logMAR | -0.352 | -1.355 | 0.651 | 0.491 |
| IOP, mmHg | 0.048 | -0.001 | 0.097 | 0.055 |
| CCT, μm | 0.001 | -0.003 | 0.006 | 0.640 |
| Axial length, mm | 0.037 | -0.117 | 0.190 | 0.640 |
| Cornea diameter, mm | -0.222 | -0.548 | 0.103 | 0.180 |
| Choroidal thickness, μm | 0.001 | -0.001 | 0.003 | 0.407 |
| Retinal thickness, μm | -0.0003 | -0.008 | 0.007 | 0.945 |
| DR status |  |  |  |  |
| Non DR | 1.0(ref) |  |  |  |
| Mild DR | -1.369 | -1.787 | -0.951 | **<0.001** |
| VTDR | -1.071 | -1.607 | -0.536 | **<0.001** |

DR=diabetic retinopathy; VTDR= vision threatening diabetic retinopathy; BCVA=best corrected visual acuity; IOP= intraocular pressure; CCT=central corneal thickness. Bold indicates statistically significant.

**Table 3.** Stepwise multivariable linear regression analysis for relationship between average parafoveal vessel density and diabetic retinopathy.

|  | Coefficient | 95%CI |  | P-value |
| --- | --- | --- | --- | --- |
|  |  | Lower | Upper |  |
| **Presence of DR** |  |  |  |  |
| Presence or non DR | -1.062 | -1.424 | -0.699 | **<0.001** |
| Age, year | -0.022 | -0.041 | -0.004 | **0.019** |
| HbA1c, % | -0.185 | -0.287 | -0.084 | **<0.001** |
| **DR severity** |  |  |  |  |
| Mild DR vs non-DR | -1.182 | -1.610 | -0.755 | **<0.001** |
| VTDR vs non-DR | -0.759 | -1.317 | -0.202 | **0.008** |
| Age, year | -0.022 | -0.040 | -0.003 | **0.022** |
| Gender (male vs female) | -0.271 | -0.539 | -0.003 | **0.048** |
| HbA1c, % | -0.192 | -0.294 | -0.090 | **<0.001** |

Bold indicates statistically significant.

**Figure 1.**
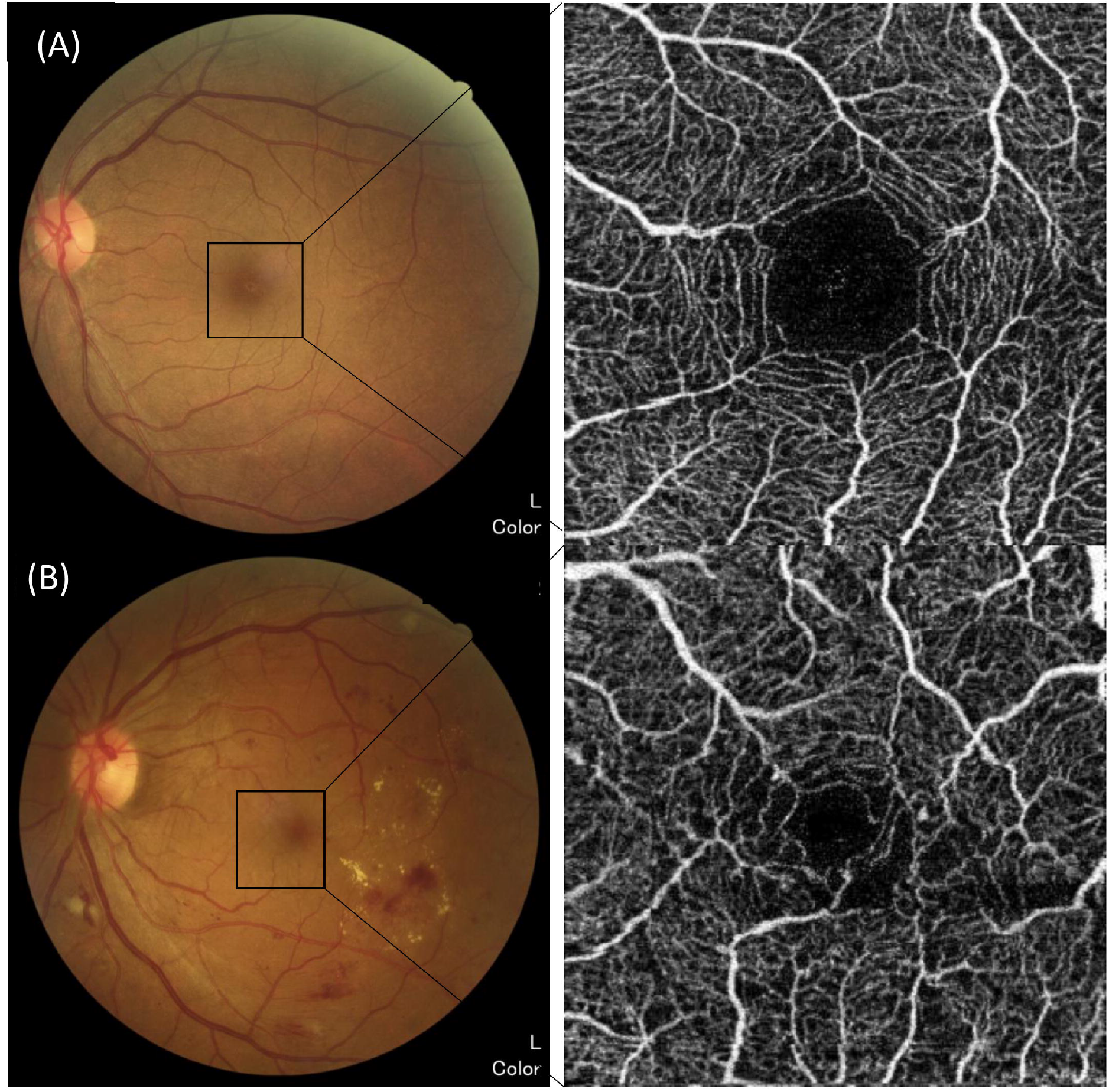
Example of fundus photography and images of optical coherence tomography angiography. (A) Diabetes mellitus without retinopathy showing intact borderline of foveal avascular zone and normal retinal capillary plexus; (B) Patients with diabetic retinopathy showing disruption of foveal avascular zone and dropouts of retinal capillary plexus.

**Figure 2.**
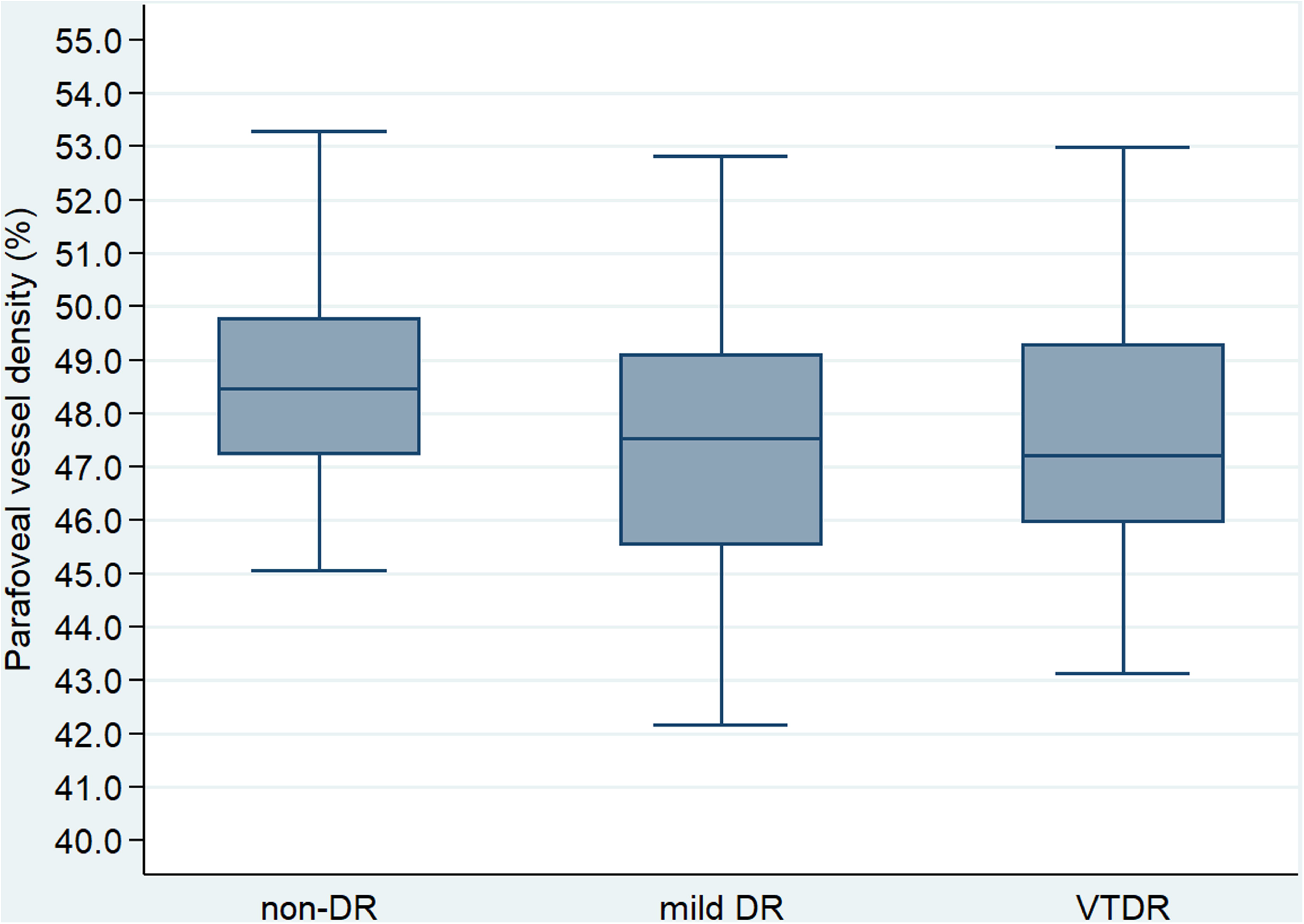
Boxplots showing the distribution of average parafoveal vessel density by status of retinopathy. DR=diabetic retinopathy; VTDR=vision threatening diabetic retinopathy.

The presence of DR served as the independent variable in the first step, and the results showed a negative correlation between PVD and the presence of DR (β = −1.062, 95%CI = −1.424 to −0.699, P < 0.001), age (β=−0.022, 95%CI = −0.041 to −0.004, P = 0.019), and HbA1c (β= −0.185, 95%CI = −0.287 to −0.084, P < 0.001). When taking DR severity as the independent variable in the second step, there was a negative correlation between PVD and DR severity, age, gender, and HbA1c.

The association of PVD in different quadrants and DR was also analysed, as shown in Table 4. Patients with DR had lower PVD in superior, inferior, and temporal quadrants (after adjusting for age, gender and HbA1c) than those without DR. Similar results were shown when comparing the mild DR group and non-DR group. Regarding the VTDR group, only the PVD in the superior quadrant was significantly less than that of patients without DR.

**Table 4.** Association between parafoveal vessel density in each quadrant and diabetic retinopathy.

|  | Univariate |  |  | Multivariate* |  |  |
| --- | --- | --- | --- | --- | --- | --- |
| | $\beta$ | 95%CI | P | $\beta$ | 95%CI | P |
| **Any DR vs non-DR** |  |  |  |  |  |  |
| Superior | -1.723 | (-2.506, -0.940) | **<0.001** | -1.409 | (-2.235, -0.583) | **0.001** |
| Inferior | -1.744 | (-2.523, -0.966) | **<0.001** | -1.512 | (-2.334, -0.690) | **<0.001** |
| Nasal | -0.228 | (-0.791, 0.336) | 0.428 | -0.062 | (-0.657, 0.534) | 0.839 |
| Temporal | -1.346 | (-1.963, -0.728) | **<0.001** | -1.105 | (-1.754, -0.455) | **0.001** |
| **Mild DR vs non-DR** |  |  |  |  |  |  |
| Superior | -1.643 | (-2.595, -0.692) | **0.001** | -1.372 | (-2.346, -0.398) | **0.006** |
| Inferior | -1.924 | (-2.869, -0.978) | **<0.001** | -1.725 | (-2.694, -0.755) | **0.001** |
| Nasal | -0.420 | (-1.105, 0.264) | 0.228 | -0.265 | (-0.967, 0.436) | 0.458 |
| Temporal | -1.489 | (-2.239, -0.739) | **<0.001** | -1.272 | (-2.038, -0.506) | **0.001** |
| **VTDR vs non-DR** |  |  |  |  |  |  |
| Superior | -1.861 | (-3.081, -0.641) | **0.003** | -1.478 | (-2.749, -0.208) | **0.023** |
| Inferior | -1.434 | (-2.646, -0.222) | **0.020** | -1.113 | (-2.378, 0.151) | 0.084 |
| Nasal | 0.106 | (-0.771, 0.984) | 0.812 | 0.319 | (-0.596, 1.235) | 0.494 |
| Temporal | -1.097 | (-2.059, -0.135) | **0.025** | -0.790 | (-1.789, 0.209) | 0.121 |

\*Adjusted for age, gender, HbA1c.
DR=diabetic retinopathy; VTDR= vision threatening diabetic retinopathy.
Bold indicates statistically significant.

### Association between PVD and other parameters

Univariable linear regression analysis revealed a negative correlation between PVD and age (P = 0.049), gender (P = 0.020), duration of diabetes (P = 0.039), use of insulin (P = 0.025), and HbA1c (P < 0.001), as shown in Figure 3. There was no significant association between PVD and axial length or other parameters. When applying multivariable linear regression, age and HbA1c maintained a negative correlation with PVD, as shown in Table 3.

**Figure 3.**
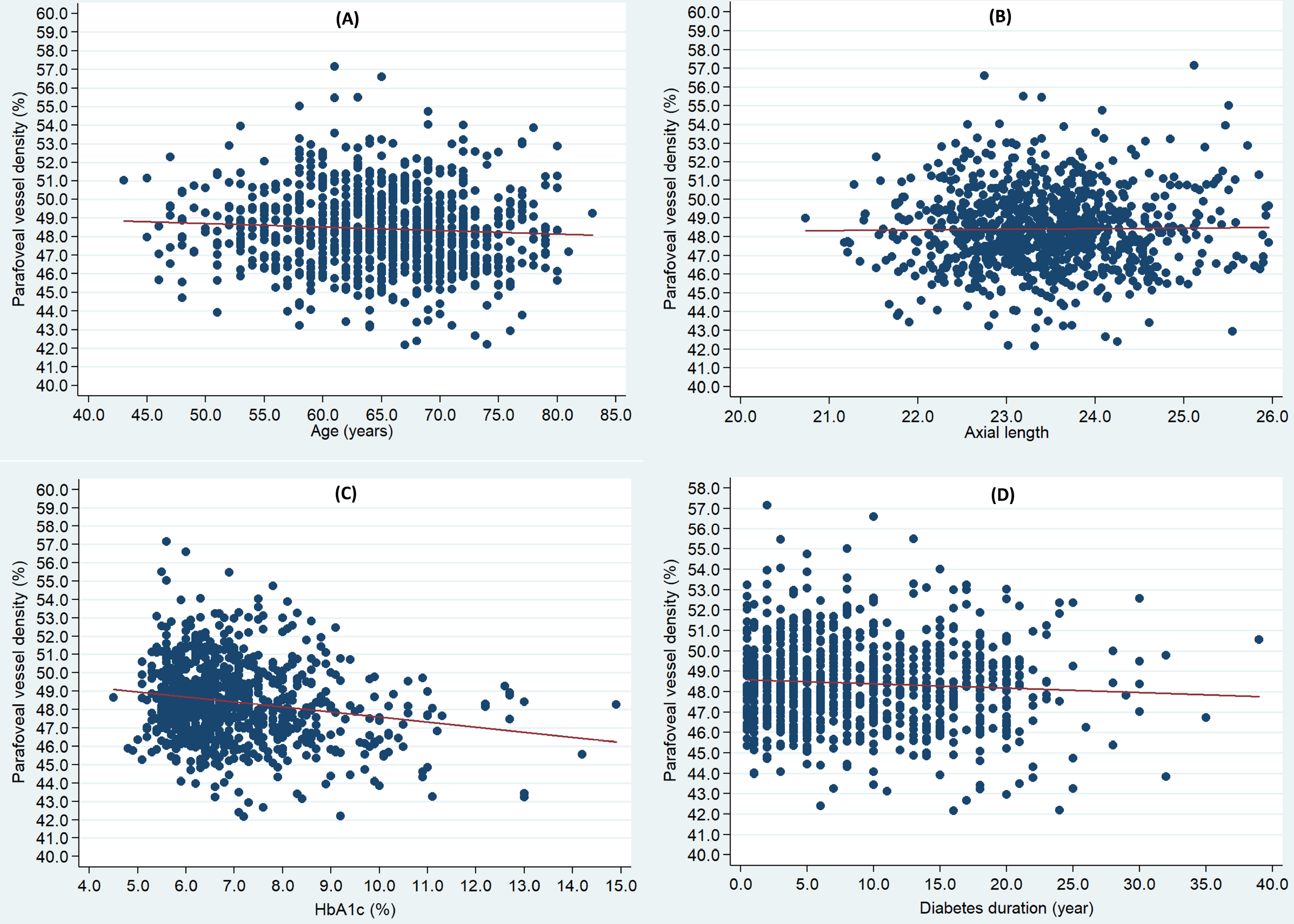
Scatter plots showing the relationship between average parafoveal vessel density and clinical factors. (A) average parafoveal vessel density versus age; (B) average parafoveal vessel density versus axial length; (C) average parafoveal vessel density versus HbA1C level; (D) average parafoveal vessel density versus duration of diabetes mellitus.

The association between PVD and clinically significant diabetic macular edema (DME) was explored (Table 5). Participants with DME had a significantly lower average PVD and temporal PVD than those without DME when adjusted for confounding factors.

**Table 5.** Association between parafoveal vessel density and clinically significant diabetic macular edema.

| DME vs no DME | Coefficient | 95%CI |  | P-value |
| --- | --- | --- | --- | --- |
|  |  | Lower | Upper |  |
| **Univariable model** |  |  |  |  |
| Average PVD | -0.907 | -1.707 | -0.108 | **0.026** |
| Superior PVD | -0.624 | -2.417 | 1.170 | 0.495 |
| Inferior PVD | -0.651 | -2.434 | 1.131 | 0.474 |
| Nasal PVD | -0.130 | -1.409 | 1.149 | 0.842 |
| Temporal PVD | -2.224 | -3.631 | -0.817 | **0.002** |
| **Age and sex adjusted** |  |  |  |  |
| Average PVD | -0.844 | -1.642 | -0.046 | **0.038** |
| Superior PVD | -0.524 | -2.319 | 1.270 | 0.566 |
| Inferior PVD | -0.652 | -2.439 | 1.136 | 0.474 |
| Nasal PVD | -0.073 | -1.354 | 1.207 | 0.911 |
| Temporal PVD | -2.128 | -3.531 | -0.724 | **0.003** |
| **Age, sex, HbA1c adjusted** |  |  |  |  |
| Average PVD | -0.790 | -1.575 | -0.005 | **0.049** |
| Superior PVD | -0.453 | -2.238 | 1.333 | 0.619 |
| Inferior PVD | -0.582 | -2.361 | 1.197 | 0.521 |
| Nasal PVD | -0.047 | -1.326 | 1.233 | 0.943 |
| Temporal PVD | -2.078 | -3.477 | -0.680 | **0.004** |

PVD=Parafoveal vessel density; DME=Diabetic macular edema.
Bold indicates statistically significant.

## Discussion

To the best of our knowledge, this is the first study that use SS-OCTA to investigate the association of macular VD and DR in diabetic patients from Chinese communities. The results demonstrated that PVD was significantly reduced in DR patients, and that it was negatively correlated with the stage of DR, especially in the superior, inferior and temporal quadrants. PVD was correlated with age and HbA1c and DME, but not axial length. The findings support a close relationship between the alterations of retinal microcirculation and the pathogenesis of DR.

The relationship between VD and DR is controversial. Forte et al.[19] reported decreased VD in non-DR and mild DR patients, which indicates that the papillary loses or lacks perfusion in the early stages of the disease. Dimitrova et al.[20] reported that the superficial and deep PVD in diabetic patients without DR decreased, when compared with healthy participants. Li et al.[21] analysed the VD of 97 DM patients and 48 controls and found that VD decreased in the diabetic patient group. However, Al-Sheikh et al.[22] found that VD increased in the DR group, when compared to those without DR. This study found that PVD changes in patients were associated with the presence of DR and the DR severity, providing new and reliable evidence for confirming the relationship between VD and DR.

There are several reasons that may account for the decrease of VD in patients with DR. Firstly, the sustained hyperglycaemia environment promotes the damage of blood-retina barrier, the proliferation of microvascular endothelial cells (leading to the narrowing or occlusion of the capillary canal), and the final atrophy and degeneration.[23] Secondly, the enlargement of endothelial cell space in retinal microvasculature is related to the stimulation of inflammatory responses, which produces exudate that accumulates in the tissues. This can compress peripheral capillaries and affect tissue metabolism, further aggravating capillary occlusion and degeneration.[24] Lastly, it could also be due to the strong oxygen consumption and metabolism of retinal photoreceptors, which produce a large amount of oxidative stress and directly stimulate the surrounding tissue structure.[25, 26] This has a greater impact in the deep retinal microvascular tissue. Further experimental studies are needed to explain the underlying mechanisms.

Few studies have analysed the association between VD and blood glucose. Inanc et al.[17] found that the course of DM disease and HbA1c level were not correlated with OCTA parameters in 60 DM patients without DR and 57 age-matched controls. Cao et al.[27] also found that the mean VD of superior and deep capillary plexus was independent of HbA1c in DM patients. Bhanushali et al.[15] reported that the spacing of large blood vessels in the superficial retinal layer was positively correlated with HbA1c, fasting blood glucose values and postprandial blood glucose values in patients with type 2 diabetes. In a Portuguese study, it was reported that vascular density, FAZ and other OCTA indicators were not associated with HbA1c or the duration of the disease.[28] When using OCTA and flicker electroretinography (ERG) examinations, Zeng et al.[29] found that the blood flow density of the superficial and deep retinal capillary networks increased in patients with type 2 diabetes patients without DR. It also showed that the increase of HbA1c and decrease of blood flow density were correlated with degree of ERG abnormalities. Our study found that HbA1C was independently correlated with retinal blood flow density based on a large-scale sample.

The relationship between VD and DME has been disputed. Hsieh et al. found that the baseline superficial PVD in DME patients predicted vision after ranibizumab treatment.[30] However, Busch et al.[31] did not observe any changes of OCTA parameters before and after intravitreal aflibercept. Hwang et al.[32] compared 12 cases of DR with 12 healthy individuals, and reported that the VD in the DR group decreased and the avascular zone increased. Kim et al. found that the VD in patients with severe NPDR and proliferative DR was significantly lower than that in patients with mild NPDR and normal controls.[14] They found that patients with DME had significantly lower VD. This study demonstrated that DME had a significant negative correlation with average PVD and temporal PVD.

VD is closely related to the range of nonperfusion vessels. The early stages of damage to the retinal capillaries will not form a region without local perfusion, but will show a reduction of VD.[33] Therefore, VD changes can also be one of the specific indicators of early DR.[34] Fluorescein angiography (FA) is the main clinical observation method for retinal microcirculation, but it is limited: the retinal vascular images provided are not complete and sometimes lack image information for the deep capillary network. OCTA, in comparison to FA, is not affected by the metabolism of fluorescent agents and image superposition can be carried out to improve the resolution of the image. The acquired retinal vascular perfusion image can make up for part of the imaging deficiency of FA.[32, 35]

This study has some advantages. Firstly, confounding factors, such as age, gender, blood pressure, blood lipids, PRP, and anti-VEGF, were not adequately corrected in previous studies.[36] The present study included patients who were unaware of ocular treatment. It was also adjusted for the confounding factors, rendering the results more reliable and accurate. Secondly, this study had a large sample size and automatically quantified parameters, making the results objective and accurate. Thirdly, all participants were homogenous DM patients from the same Chinese communities. There are still some limitations in this study. Firstly, the blood-retina barrier function could not be evaluated by OCTA technology, which is important phenotype of DR.[37] However, it was not feasible to perform FFA for this large sample size of relative normal DM patients. Secondly, the range of observation of this study is 3 mm × 3 mm, but the pathology alteration of DR could exist in any region of the retina. Thus, a larger scan area is needed in future studies. However, a recent study reported that 3×3 mm OCTA images detected DR best.[38] Third, the nature of the cross-sectional design prevents cause-effect inference in macular ischemia and DR development. Further longitudinal group studies are needed.

In summary, this study demonstrated that PVD decreased in patients with DR. Age, HbA1c and DME were also negatively correlated with PVD. These findings provide more evidence to suggest that microcirculation alterations play a role in the pathogenesis of DR. Further studies are needed to examine the possible mechanism behind the findings of this study.

## Data Availability

Available when appropriate request.

## Acknowledgements

None.

## Author contributions

Design and conduct of the study (WW, NX, WH); Collection, management, analysis and interpretation of the data (NX, YX, SS, WW, WH); Preparation of the manuscript (NX, YT, SL, WW, WH); Review and final approval of the manuscript (all authors).

## Compliance with ethical standards

### Human and animal rights

This article does not contain any studies with human participants or animals performed by any of the authors.

### Ethical approval

The study was approved by the ethnic committee of ZOC.

### Informed consent

For this type of study, informed consent was not required.

### Data availability statements

The datasets generated during and/or analyzed during the current study are available from the corresponding author on reasonable request.

